# Prognostic Value of Plasma Glial Fibrillary Acidic Protein in Cognitively Unimpaired Older Adults: Results from the A4 Study

**DOI:** 10.1101/2025.08.28.25334681

**Authors:** Elham Ghanbarian, Tianchen Qian, Babak Khorsand, Lukai Zheng, S. Ahmad Sajjadi, Joshua D. Grill, Laura A. Rabin, Richard B. Lipton, Reisa A. Sperling, Ali Ezzati

**Author notes:** **Corresponding author:** Elham Ghanbarian, MD, Ph.D.

## Abstract

**INTRODUCTION:** Plasma glial fibrillary acidic protein (GFAP), a marker of astrocytic activation, has been linked to Alzheimer’s disease; however, its prognostic value in cognitively unimpaired (CU) individuals remains unclear.

**METHODS:** We included 949 CU older adults from the A4 preclinical AD trial, and its companion LEARN cohort. Baseline plasma GFAP was measured, and longitudinal associations with cognitive decline, clinical dementia rating (CDR) progression, and imaging biomarkers were assessed over 240 weeks.

**RESULTS:** Baseline plasma GFAP was higher in females and in A4 (amyloid-positive) versus LEARN (amyloid-negative) participants. Cross-sectionally, elevated GFAP was associated with lower cognitive performance and greater amyloid burden. Longitudinally, higher GFAP predicted faster cognitive decline, increased risk of CDR progression, AD-related cortical atrophy, and amyloid conversion, with stronger effects in females.

**DISCUSSION:** Plasma GFAP is a prognostic biomarker in CU older adults, predicting cognitive and biological changes, with stronger associations observed in females, highlighting a possible sex-specific vulnerability.

## 1. Introduction

Pathological processes in Alzheimer’s disease (AD) begin decades before symptom onset,^1^ making the preclinical phase a critical window for risk stratification and intervention. Blood-based biomarkers are particularly attractive for population-level screening because they are scalable, less invasive, and lower cost than PET or CSF, but their prognostic performance in cognitively unimpaired (CU) older adults remains incompletely defined.

Glial fibrillary acidic protein (GFAP) is a marker of astrocytic activation that has been linked to amyloid β pathology and cognitive decline.^2, 3^ GFAP reflects astrocytic responses to neuronal injury, a process commonly observed in proximity to amyloid plaques in patients with mild cognitive impairment (MCI) and AD.^4, 5^ Elevated GFAP has been reported in the CSF of patients with AD and other dementias,^6^ and recent studies have demonstrated higher plasma GFAP in AD dementia compared to controls.^7–9^ Plasma GFAP may help differentiate AD from other dementias and distinguish clinical stages across the AD continuum.^7, 8, 10–12^ In patients with MCI, it can detect underlying pathology, predict cognitive decline, and forecast conversion to AD dementia.^13, 14^ It also correlates with other AD biomarkers including amyloid and tau burden, as well as cortical atrophy.^15, 16^ However, its prognostic value in CU individuals is less well defined. Existing studies suggest that associations between GFAP, cognition, and AD biomarkers are often limited to amyloid-positive individuals,^17–20^ highlighting the need to consider amyloid status as a potential modifier.

Sex and age may influence the relationship between GFAP and disease outcomes. Women have a higher lifetime risk of AD and often show greater neuropathologic burden, which may be partly explained by genetic, hormonal, and immunologic factors. Evidence from both animal and human studies suggests that females mount stronger innate immune responses,^21^ which may amplify neuroinflammation reflected by GFAP. While heightened immune activity has been linked to the higher prevalence of AD in females, it remains unclear whether GFAP provides greater prognostic value in this population. Age is another critical modifier and while midlife inflammation is generally associated with increased dementia risk, in the oldest-old the relationship may weaken or even reverse.^22–24^ Understanding whether GFAP’s prognostic associations differ by sex and age is therefore important, as these demographic factors may alter the biological pathways linking astrocytic activation to cognitive decline and neurodegeneration.

In this study, we examined the association of baseline plasma GFAP with the rate of cognitive decline, risk of clinical progression, and longitudinal changes in structural MRI, amyloid PET, and plasma p-tau217 in CU participants from the Anti-Amyloid Treatment in Asymptomatic Alzheimer’s Disease (A4) Study. We further investigated whether these associations differ by amyloid status, sex, and age to better understand the potential of plasma GFAP as an early marker of disease risk across diverse subgroups.

## 2. Methods

### 2.1. Participants and study design

#### 2.1.1. A4 and LEARN study design

We used data from the Anti-Amyloid Treatment in Asymptomatic Alzheimer’s Disease (A4) and its observational companion, the longitudinal Evaluation of Amyloid Risk and Neurodegeneration (LEARN) studies. Detailed methodology for A4 and LEARN screening has been described previously. ^25, 26^ Briefly, the A4 study was a 240-week, multicenter, double-blind, placebo-controlled randomized clinical trial designed to evaluate whether solanezumab slows cognitive decline in preclinical AD. A4 enrolled 1169 healthy, CU adults aged 65 to 85, with Clinical Dementia Rating (CDR) score of 0, Mini-Mental State Examination (MMSE) of 25-30, and Wechsler Memory Scale-Revised, Logical Memory IIa Delayed Recall score of 6-18, who were defined as amyloid-positive via PET imaging as described below. Participants with elevated amyloid on screening amyloid PET and who met the inclusion and exclusion criteria were randomly assigned to either placebo (n= 583) or solanezumab (n= 564) groups. All participants received an annual CDR and cognitive testing every six months with the Preclinical Alzheimer Cognitive Composite (PACC) over the course of 4.5 years.

The LEARN study served as the amyloid-negative comparison arm. LEARN enrolled 538 CU individuals who met the identical screening criteria to A4, except that they were classified as amyloid-negative by PET. Participants in LEARN underwent the same cognitive and functional assessments as those in A4, providing a parallel longitudinal cohort of amyloid-negative older adults.

#### 2.1.2. Current study participants

The inclusion criteria and number of participants in our study are summarized in Fig 1. For the cross-sectional analysis, we included all participants from the A4 study (n = 499) who completed their baseline visit and had a baseline serum GFAP measurement, regardless of treatment group, as well as all LEARN participants with available baseline GFAP data (n = 450), for a total of 949 participants (578 females, 371 males).

**Figure 1.**
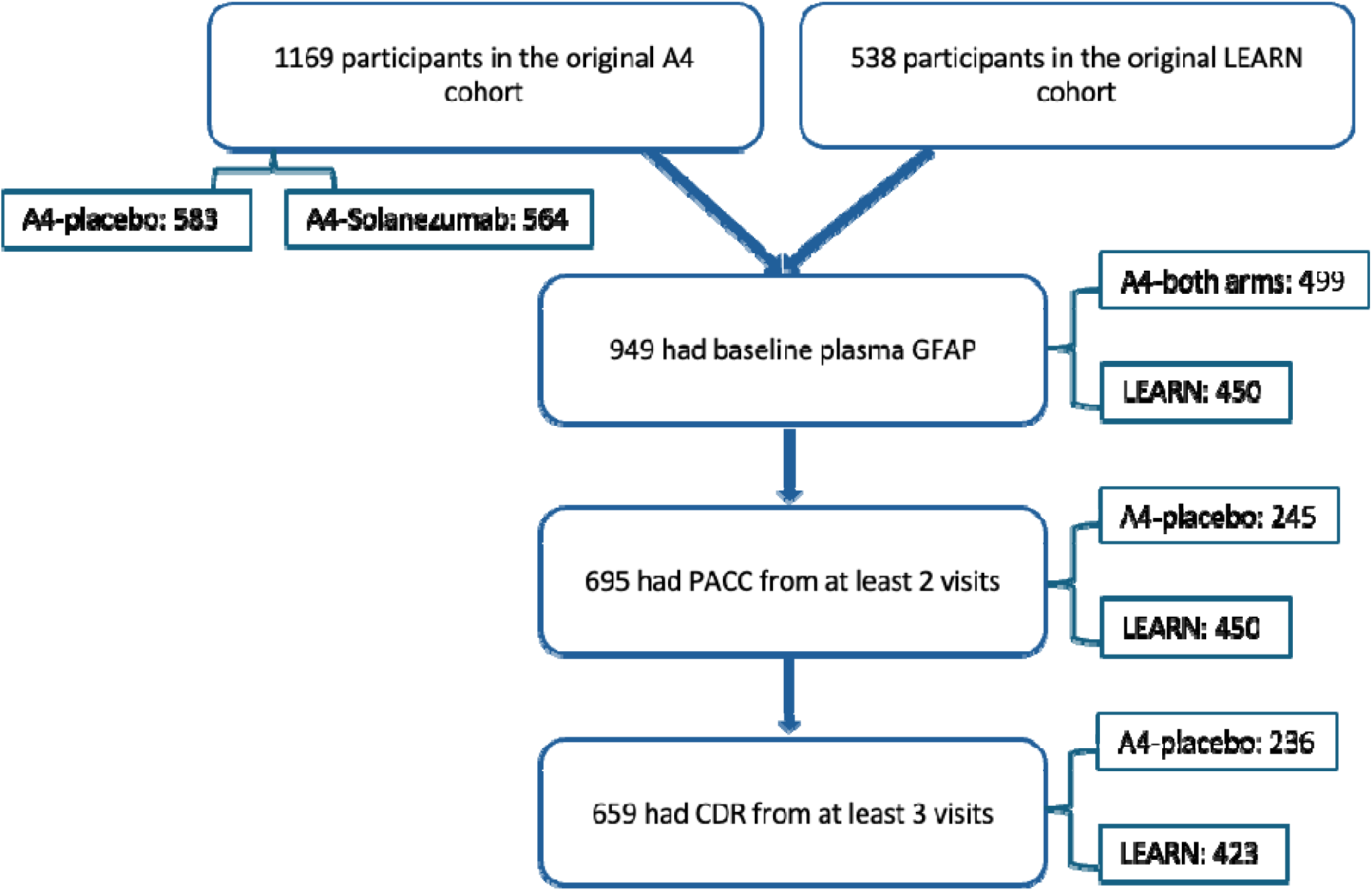
Participant flow diagram showing inclusion criteria and sample sizes for analyses.

For the longitudinal analysis, we restricted the sample to the placebo arm of the A4 study and to LEARN participants with available baseline plasma GFAP levels, excluding solanezumab-treated individuals to eliminate any potential confounding effects of the drug on cognitive and biomarker trajectories. To evaluate the association of baseline GFAP with change in cognition, participants were required to have at least two PACC assessments (n = 695; 418 females, 277 males), drawn from the A4 placebo arm (n = 245) and LEARN (n = 450).

For the analysis of clinical progression, participants with baseline GFAP and at least three global CDR ratings were included (n = 659; 392 females, 267 males), consisting of 236 A4 placebo participants and 423 LEARN participants.

For biomarker analyses, participants were included if they had both baseline plasma GFAP and the relevant biomarker (MRI, amyloid PET, or p-tau217). Longitudinal analyses required paired baseline and final visit (week 240) data. The number of participants included in each analysis is summarized in Supplementary Table 1.

### 2.2. Outcome Assessments

#### 2.2.1 Neuropsychological assessments

##### Preclinical Alzheimer’s Cognitive Composite (PACC)

The primary outcome in the A4 and LEARN studies was the Preclinical Alzheimer’s Cognitive Composite Scale (PACC).^27, 28^ PACC includes 4 components: MMSE score (0-30), digit symbol substitution test (correct responses in 90 seconds; maximum raw score = 91), LMDR-IIa score (0-25), and the free and cued selective reminding test (FCSRT; sum of free and total cued recall; score, 0-96). Each component score was standardized (z-score) using the baseline mean and standard deviation, and the PACC was calculated as the sum of the four z-scores. Negative values indicated cognitive decline. The PACC was administered at baseline and every 24 weeks thereafter.

##### The Clinical Dementia Rating (CDR)

The global CDR is a two-part structured interview conducted by a trained and certified clinician at each site.^29^ The CDR begins with a clinician interview of the study partner, and the second part involves the clinician interview of the participant. Scoring is tabulated based on the study partner interview and the clinician’s objective assessment of the participant. The global CDR is an ordinal scale ranging from 0 to 3 as follows: 0= no cognitive impairment, 0.5= mild cognitive impairment, 1= mild dementia, 2= moderate dementia and 3= severe dementia. In the A4 and LEARN studies, progression was defined as conversion from a global CDR = 0 to > 0, confirmed by either two consecutive non-zero ratings or a non-zero rating at the final visit. Time to progression was calculated as the interval between baseline and the first qualifying non-zero score, with censoring at the last available assessment for non-progressors.

#### 2.2.2 Plasma Biomarkers

##### Glial fibrillary acidic protein (GFAP)

Plasma GFAP levels were measured using the fully automated plasma NeuroToolKit (NTK), a panel of exploratory robust prototype assays (Roche Diagnostics International Ltd, Rotkreuz, Switzerland).^30^

##### Phosphorylated tau (p-tau217)

Quantification of p-tau217 was assayed on an analytically validated electrochemiluminescent (ECL) immunoassay using a Meso Scale Discovery (MSD) Sector S Imager 600 instrument at the Lilly Clinical Diagnostics Laboratory. Measurements were available from baseline and week 240 (final visit) for participants in both A4 and LEARN.^31^

#### 2.2.3 Imaging biomarkers

Volumetric MRI data were available for a subset of participants at both baseline and the final visit. Regions of interest included bilateral hippocampal volume, total cortical gray matter volume, and cortical gray matter volume in AD-signature regions (entorhinal cortex, middle temporal gyrus, inferior temporal gyrus, and fusiform gyrus). Volumes were normalized to intracranial volume to adjust for head size differences.

Brain amyloid levels were assessed using 18F-florbetapir PET imaging, with the use of a mean cortical standardized uptake value ratio (SUVR) with a cerebellar reference region. A threshold SUVR ≥ 1.15 was classified as amyloid positive. Values between 1.10–1.15 were considered elevated only if the visual read was positive. Amyloid PET was done at baseline and the visit at 240 weeks.^25^

### 2.3. Statistical Analysis

#### Group comparisons

Differences in participant characteristics across sex and age subgroups were tested using independent-sample t tests.

#### Cognitive trajectories (PACC)

To evaluate the effect of baseline plasma GFAP on baseline and longitudinal cognitive performance, we used linear mixed-effects (LME) models. Models included fixed effects for time, GFAP, and GFAP × time interaction, and covariates (age, sex, education). Random intercepts and slopes for time were included at the participant level to capture individual variability in baseline cognition and rate of cognitive change. The estimate for GFAP reflects its association with baseline PACC, while the estimate for GFAP × time interaction represents how GFAP modifies the rate of change in PACC over time.

#### Clinical progression (CDR)

Associations between baseline GFAP and progression to global CDR > 0 were examined using Cox proportional hazards models adjusted for age, sex, and education. Time was defined as weeks from baseline to the first of two consecutive nonzero CDR ratings or a nonzero rating at the final visit. Kaplan–Meier survival curves stratified by baseline GFAP quartiles were generated for visualization.

#### Cross-sectional analyses of biomarkers

To examine baseline associations between plasma GFAP and biomarkers (MRI, amyloid PET, and p-tau217), we used linear regression models adjusted for age, sex, and education.

#### Longitudinal change in biomarkers

For biomarkers with baseline and week-240 data, linear regression models adjusted for the same covariates were used to evaluate the association between baseline GFAP and biomarker change scores, defined as the difference between the final and baseline measurements.

#### Amyloid conversion (LEARN)

In the LEARN cohort, we tested whether baseline GFAP predicted incident amyloid positivity at week 240 using logistic regression models, restricted to participants who were amyloid-negative at baseline, and had follow-up PET imaging.

#### Stratified analyses

All models were repeated separately by sex, age group (65–75 vs. >75), and amyloid status (A4 vs. LEARN) to examine subgroup-specific associations. The age range of participants in the original A4 Study was 65–85 years, and in our analysis, participants were grouped into 65–75 and 75–85 years to enable comparison across two decades.

#### Multiple comparisons and software

To account for multiple comparisons, false discovery rate (FDR) correction was applied separately for baseline and longitudinal analyses. All statistical analyses were conducted in SPSS version 27.

## 3. Results

### 3.1. Cohort characteristics

Baseline GFAP data were available for 949 participants, from both arms of the A4 study (n=499) and the LEARN study (n=450). The entire sample consisted of 578 females (60.9%), and the average age across all participants was 70.94 ± 4.69 years. Males were on average older than females (71.71 ± 4.94 vs. 70.76 ± 4.38, *p* = 0.003) and had higher educational attainment (17.27 ± 2.67 vs. 16.28 ± 2.46 years, *p* < 0.001). The average plasma GFAP level in the entire sample was 0.11 ± 0.05 ng/mL and was significantly higher in females than males (0.12 ± 0.06 vs. 0.10 ± 0.05 ng/mL, *p* < 0.001). In the age-stratified sample, 773 participants were included in the 65– 75-year-old group, and 176 participants were over 75 years old. The average baseline GFAP level was significantly higher in the older group (0.15 ± 0.07 vs. 0.10 ± 0.05 ng/mL, *p* < 0.001). Additionally, the average baseline GFAP level was higher in A4 participants compared to those in the LEARN study (0.12 ± 0.06 vs. 0.09 ± 0.04 ng/mL, *p* < 0.001), with higher GFAP levels in females in both cohorts (both *p* < 0.001). Demographic, clinical, and biomarker characteristics of the total sample, as well as those stratified by sex, are summarized in Table 1.

**Table 1.**
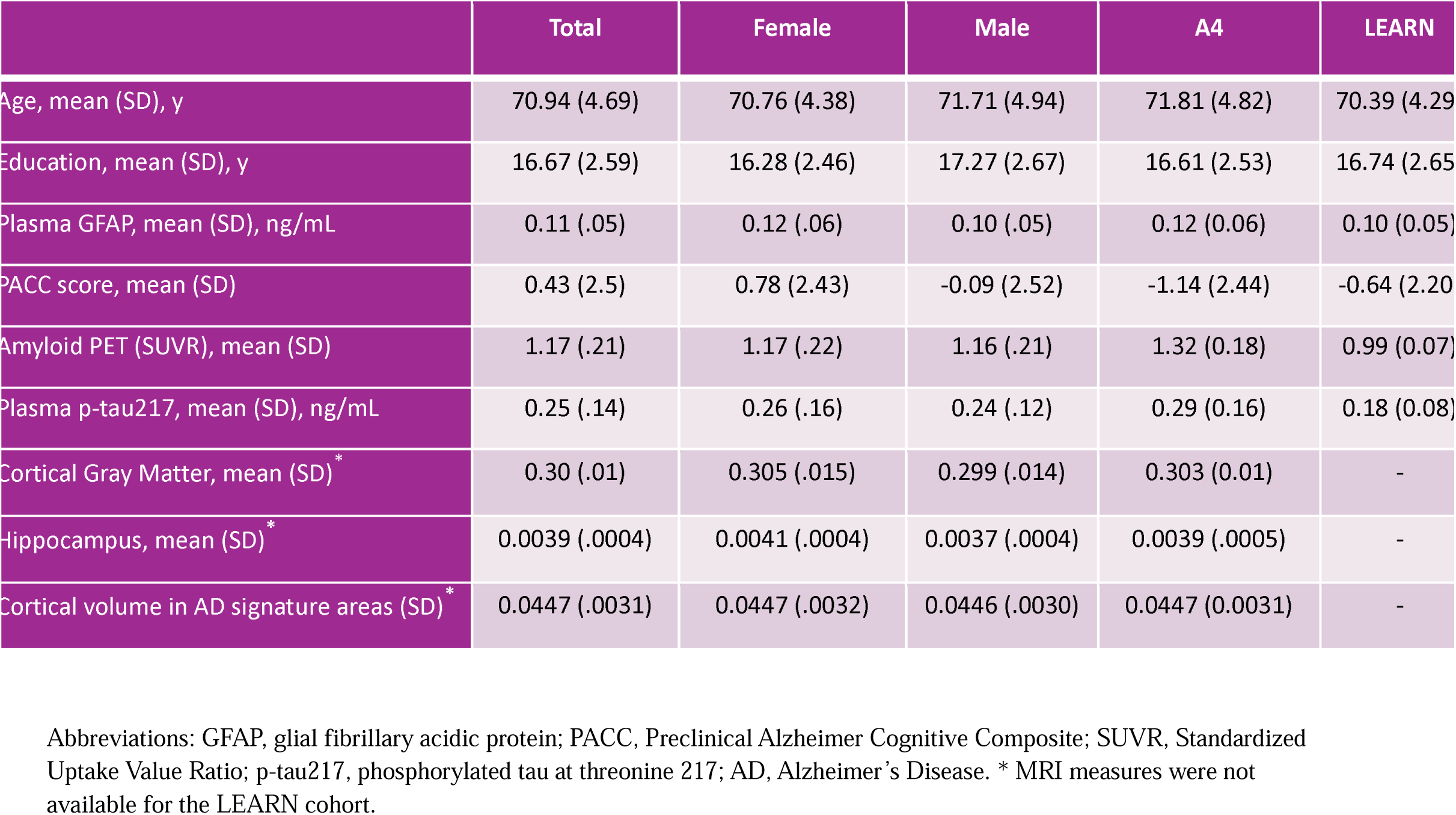
Baseline demographic, cognitive, and biomarker characteristics of participants by sex and cohort.

### 3.2. Association of baseline GFAP with cognitive performance (PACC)

We investigated the associations between baseline plasma GFAP levels and trajectories of cognitive decline as measured by PACC scores (Table 2). In the overall sample, higher baseline GFAP levels were associated with lower baseline PACC scores (β = -4.34 ± 1.52, *p* = 0.008) and a faster rate of decline in PACC over time (β = -0.17 ± 0.03, *p* < 0.001). In the sex-stratified analyses, GFAP was significantly associated with both lower baseline PACC (β = -5.31 ± 1.93, *p* = 0.008) and faster decline (β = -0.21 ± 0.04, *p* < 0.001) in females. In males, GFAP was associated only with the rate of decline (β = -0.14 ± 0.06, *p* = 0.018), but not with the baseline performance. In the age-stratified analyses, GFAP was associated with both baseline PACC and rate of change in both age groups. The results of the age- and sex-stratified analyses are presented in Supplementary Tables S2 and S3.

**Table 2.** Associations of baseline plasma GFAP with baseline performance and longitudinal change in PACC.

| cohort | Total (A4+LEARN) |  |  |  |  |  | A4 |  |  |  |  |  | LEARN |  |  |  |  |  |
| --- | --- | --- | --- | --- | --- | --- | --- | --- | --- | --- | --- | --- | --- | --- | --- | --- | --- | --- |
|  | Baseline PACC |  |  | Rate of change |  |  | Baseline PACC |  |  | Rate of change |  |  | Baseline PACC |  |  | Rate of change |  |  |
|  | β | SE | p value* | β | SE | p value* | β | SE | p value* | β | SE | p value* | β | SE | p value* | β | SE | p value* |
| entire cohort | -4.34 | 1.52 | 0.008 | -0.17 | 0.03 | <0.001 | -2.22 | 2.26 | 0.612 | -0.20 | 0.06 | 0.010 | -6.27 | 2.17 | 0.007 | -0.02 | 0.03 | 0.8 |
| female | -5.31 | 1.93 | 0.008 | -0.21 | 0.04 | <0.001 | -2.40 | 3.07 | 0.612 | -0.20 | 0.09 | 0.045 | -8.62 | 2.60 | 0.003 | -0.03 | 0.04 | 0.8 |
| male | -2.67 | 2.48 | 0.283 | -0.14 | 0.06 | 0.018 | -2.07 | 3.35 | 0.612 | -0.17 | 0.10 | 0.106 | -0.74 | 3.84 | 0.848 | -0.05 | 0.07 | 0.8 |
| age 65-75 | -5.80 | 1.89 | 0.008 | -0.10 | 0.04 | 0.018 | -1.48 | 2.92 | 0.612 | -0.14 | 0.09 | 0.106 | -8.37 | 2.46 | <0.001 | -0.01 | 0.04 | 0.8 |
| age >75 | -7.49 | 2.68 | 0.008 | -0.21 | 0.08 | 0.017 | -7.78 | 3.44 | 0.140 | -0.23 | 0.10 | 0.045 | -4.77 | 4.26 | 0.333 | -0.05 | 0.01 | 0.8 |

**Total (A4 + LEARN):** n=695; Female n=418, Male n=277, Age 65-75 n= 573, Age>75 n= 122; **A4:** n=245; Female n=140, Male n=105, Age 65-75 n= 182, Age>75 n= 63; **LEARN:** n=450; Female n=278, Male n=172, Age 65-75 n= 391, Age>75 n= 59. \*All p values are FDR adjusted.

In the analysis stratified by amyloid status (A4 vs. LEARN), higher baseline GFAP levels were associated with a faster rate of decline in PACC scores among A4 participants (β = -0.20 ± 0.06, *p* = 0.010). This effect remained significant only in females (β = -0.20 ± 0.09, *p* = 0.045), and in older age (β = -0.23 ± 0.1, *p* = 0.045). In contrast, among LEARN participants, higher GFAP levels were associated with lower baseline PACC scores (β = -6.27 ± 2.17, *p* = 0.007), but not with the rate of change over time. In the sex-stratified analysis, this association remained significant only in females (β = -8.62 ± 2.60, *p* = 0.003), and younger age (β = -8.37 ± 2.46, *p* < 0.001).

### 3.3. Association of baseline GFAP with risk of clinical progression

We examined the association between baseline plasma GFAP and risk of clinical progression, defined as progression to CDR> 0 over two follow-up visits (Table 3). In the total sample, higher baseline GFAP levels were significantly associated with increased risk of progression (HR = 1.24, 95% CI: 1.08-1.41, *p* = 0.01). In the sex stratified analysis, this association was significant in females (HR = 1.34, 95% CI: 1.1-1.62, *p* = 0.007, Fig 2A.) but not in males. The age-stratified analysis revealed that the association remained significant in both the 65–75 (n = 545, HR = 1.31, 95% CI: 1.08-1.59, *p* = 0.01) and >75 age groups (HR = 1.26, 95% CI: 1.05-1.51, *p* = 0.019). In the age- and sex-stratified analyses, significant associations were observed among females in both age groups, while no associations were found in males (Supplementary Table S4). In the amyloid-stratified analysis, higher GFAP levels were significantly associated with the increased risk of progression among amyloid negative participants from the LEARN study (HR = 1.45, 95% CI: 1.12-1.86, *p* = 0.02). In contrast, no significant associations were observed among amyloid-positive participants from the A4 study (Table 3 and Figure 2B).

**Table 3.** Associations between baseline GFAP and risk of clinical progression (CDR > 0)

| Cohort | Total (A4+LEARN) |  | A4 |  | LEARN |  |
| --- | --- | --- | --- | --- | --- | --- |
|  | Hazard Ratio | p value * | Hazard Ratio | p value * | Hazard Ratio | p value * |
| Entire cohort | 1.24 | 0.010 | 1.05 | 0.967 | 1.45 | 0.020 |
| Female | 1.34 | 0.007 | 1.14 | 0.750 | 1.43 | 0.066 |
| Male | 1.16 | 0.119 | .97 | 0.841 | 1.47 | 0.055 |
| Age 65-75 | 1.31 | 0.010 | 1.05 | 0.911 | 1.46 | 0.062 |
| Age> 75 | 1.26 | 0.019 | 1.22 | 0.440 | 1.35 | 0.129 |

**Total (A4 + LEARN):** n=659; Female n=392, Male n=267, Age 65-75 n= 545, Age>75 n= 114; **A4:** n=236; Female n=132, Male n=104, Age 65-75 n= 175, Age>75 n= 6; **LEARN:** n=423; Female n=260, Male n=163, Age 65-75 n= 370, Age>75 n= 53. \*All p values are FDR adjusted.

**Figure 2.**
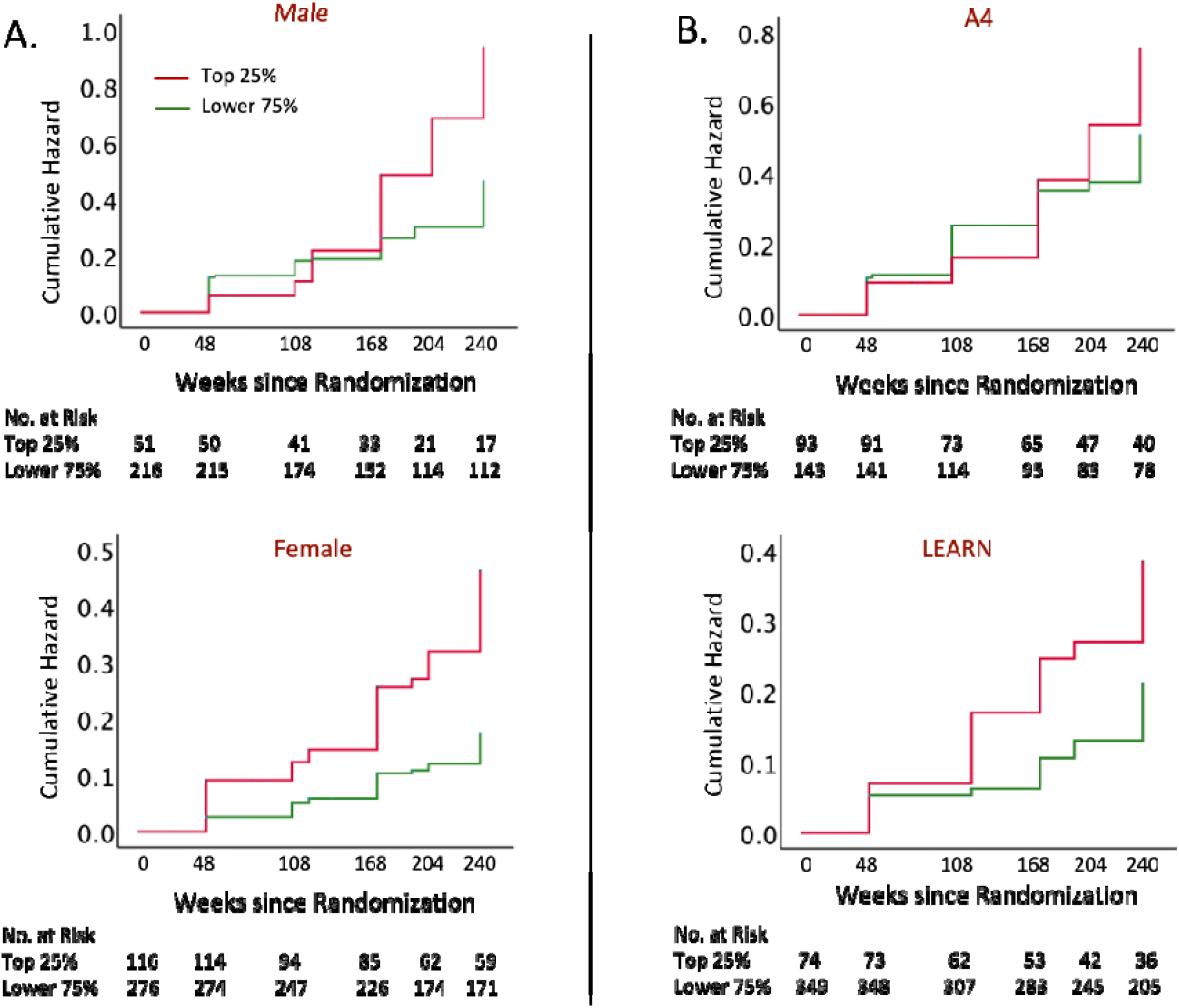
Cumulative hazard of progression to CDR > 0 stratified by baseline plasma GFAP levels, shown by sex (A) and cohort (B).

### 3.4. Association of baseline GFAP with brain atrophy

In cross-sectional analysis of the entire sample, higher baseline GFAP was associated with reduced cortical volume in AD signature regions (β = –5.95 ± 2.44, *p* = 0.015); however, this association was not significant within either sex subgroup. In a subset of participants from the placebo arm of the A4 study with available longitudinal MRI data (baseline and final visits), higher baseline GFAP was associated with greater cortical atrophy in AD signature regions over approximately five years (β = –5.51 ± 2.40, *p* = 0.023). Interestingly, in the sex-stratified analyses, this association was significant only in females (β = –9.37 ± 3.12, *p* = 0.003), and not in males.

Similarly, in the cross-sectional analysis of the same participants, there was a significant association between baseline GFAP and total cortical gray matter volume in the overall group (β = –2.78 ± 1.14, *p* = 0.015), but not within sex subgroups. In the longitudinal subsample, higher baseline GFAP predicted greater loss in total cortical gray matter volume at follow-up (β = –2.12 ± 0.95, *p* = 0.026). This association was significant in females (β = –2.97 ± 1.22, *p* = 0.017), but not in males. In both cross-sectional and longitudinal analyses, baseline GFAP was not associated with hippocampal volume in the overall sample or in sex-stratified analysis.

### 3.5. Association of GFAP with amyloid PET burden

To assess whether plasma GFAP was associated with the severity of amyloid-β deposition, we built linear regression models using the neocortical composite values of amyloid-β-PET SUVR as the outcome and plasma GFAP as the predictor. In the cross-sectional analysis of the overall sample, higher baseline GFAP was significantly associated with greater amyloid burden (β = 1.42 ± 0.13, *p* < 0.001). This association remained significant in both females and males when analyzed separately (both *p* < 0.001). In cohort-stratified analysis, in A4 participants, higher GFAP levels were associated with greater amyloid burden (β = 1.14 ± 0.13, *p* < 0.001), whereas no significant association was observed in the LEARN cohort. In longitudinal analyses, higher baseline GFAP was significantly associated with greater increase in amyloid PET over time in the overall sample (β = 0.25 ± 0.12, *p* = 0.038). This association was not significant within sex subgroups or when stratified by study cohorts.

### 3.6. Baseline GFAP and risk of conversion to amyloid positivity in LEARN

We investigated whether plasma GFAP levels could predict incident amyloid positivity at the final visit (week 240) among individuals who were amyloid-negative at baseline (defined as SUVR < 1.15). Over a follow-up period of 240 weeks, 38 participants (15.4%) converted to amyloid-positive status, including 24 females (15.7%) and 14 males (15.1%). In logistic regression models, higher baseline GFAP (z-scored) was significantly associated with increased odds of becoming amyloid-positive (OR = 1.47, 95% CI: 1.06–2.04, *p* = 0.021) in the entire population. In the sex-stratified analyses, this association remained significant in females (OR = 1.54, 95% CI: 1.01–2.36, *p* = 0.044), but not in males.

### 3.7. Association of GFAP with plasma p-tau217

In cross-sectional analysis, higher baseline GFAP levels were associated with higher p-tau217 in the entire sample (β = 1.26 ± 0.09, *p* < 0.001), as well as in females (β = 1.38 ± 0.12, *p* < 0.001) and males (β = 1.01 ± 0.13, *p* < 0.001). We also investigated the relationship of baseline GFAP with change in p-tau217 levels, defined as the difference between the final and baseline visits, in a subset of participants with available longitudinal data. No significant association was found between baseline GFAP and change in p-tau217 levels in the overall sample or in sex-stratified analyses.

## 4. Discussion

In this study of CU older adults from the A4 (Aβ+) and LEARN (Aβ-) cohorts, we found that higher baseline plasma GFAP was associated with worse baseline cognitive performance and greater amyloid burden. Longitudinally, elevated GFAP predicted faster cognitive decline, increased risk of progression to CDR > 0, greater cortical atrophy in AD-signature regions, and conversion to amyloid positivity. These associations were consistently stronger in females, who also had higher baseline GFAP levels.

Although growing evidence supports a causal role for inflammation in AD, the prognostic value of blood-based inflammatory markers in CU individuals is still not well established. Our findings indicate that plasma GFAP may serve as a prognostic marker, predicting cognitive, clinical, and imaging outcomes.

The predictive role of plasma GFAP for cognitive decline has been reported in recent studies. In longitudinal studies of CU individuals, higher plasma GFAP was associated with a steeper rate of decline in global cognition as measured by MMSE.^19, 32^ Our work extends this literature by demonstrating associations with both baseline scores and rate of decline on the PACC, a composite specifically designed to capture subtle preclinical changes.^27, 28^ This suggests that plasma GFAP is sensitive to early, preclinical cognitive decline, supporting its potential utility as a prognostic biomarker at this stage of disease.

Notably, stratified analyses showed that GFAP’s association with cognitive decline was significant only in Aβ+ individuals, whereas its association with baseline cognitive status was significant in Aβ-participants. These findings suggest that neuroinflammation, as captured by plasma GFAP, may play different roles across the AD continuum influencing cognition before significant amyloid deposition, but accelerating decline after significant amyloid deposition. Supporting this, several studies have linked plasma GFAP to both amyloid burden and cognitive decline.^14, 19, 33^ In postmortem cohorts, GFAP’s associations with tau burden and cognitive decline were most prominent in Aβ+ individuals.^34^ Similarly, in autosomal dominant AD, plasma GFAP elevations followed amyloid accumulation and preceded cognitive decline, particularly in asymptomatic individuals,^35^ suggesting that astrocytic activation is triggered or amplified by amyloid pathology. By contrast, several studies suggested that the association of GFAP and cognition may be independent of amyloid pathology. In a Chinese cohort, including cognitively normal individuals, baseline plasma GFAP levels were associated with the longitudinal decline in annual MMSE scores after adjusting for Aβ levels, implying that this association was independent of amyloid burden.^36^ Similarly, baseline plasma GFAP was found to be elevated in Aβ+ individuals and to predict longitudinal Aβ deposition in both Aβ+ and cognitively unimpaired, Aβ-participants. However, the associations between plasma GFAP and cognitive decline (MMSE) were still significant after adjusting for amyloid PET burden, indicating that, in addition to being a marker of amyloid pathology, astrocytosis could have a negative impact on cognition independent of amyloid status.^19, 32^ In summary, consistent with prior reports, we found that plasma GFAP was elevated in Aβ+ compared to Aβ-participants, predicted longitudinal Aβ increases, and was prognostic for cognitive outcomes in an amyloid-dependent manner.^17–19^

Plasma GFAP has been associated with incident AD dementia, both in patients with MCI^7, 14^ and in dementia-free adults.^37^ In CU individuals, higher baseline plasma GFAP has been linked to future conversion to AD dementia, showing stronger predictive value than other plasma biomarkers,^32, 36^ and can be elevated up to 10 years before symptom onset.^38^ In our study, GFAP predicted clinical progression to MCI in CU participants, regardless of their baseline amyloid status. In stratified analyses, GFAP predicted progression only in the LEARN cohort (Aβ-at baseline), where it also predicted conversion to amyloid positivity, but not in A4. In a prior study, plasma GFAP was associated with an increased risk of progression to dementia independent of plasma amyloid levels;^32^ however, in analyses stratified by CSF amyloid status, this association was observed only in the Aβ+ group. These findings support a model where GFAP reflects different disease stages: in Aβ-individuals, it may indicate early astrocytic activation preceding amyloid, capturing earlier clinical progression to MCI, whereas in Aβ+ individuals, the higher rate of progression to MCI (as evident in Fig 2B) is more likely driven by amyloid (and tau) and may not be captured by plasma GFAP. Alternatively, in Aβ-individuals in the LEARN cohort, GFAP may be associated with MCI risk through pathways beyond amyloid, such as vascular pathologies.^39, 40^

Our findings reveal, for the first time, a significant cross-sectional and longitudinal association between plasma GFAP and cortical atrophy in AD-signature regions, indicating astrocytic activation’s role in AD-related neurodegeneration. Higher plasma GFAP was also associated with greater total gray matter loss but showed no association with hippocampal volume. Prior studies on the association of GFAP and hippocampal volume have been inconsistent: though a meta-analysis, reported no association,^41, 42^ other studies found longitudinal decreases in cortical thickness and hippocampal volume among individuals with elevated GFAP.^10, 36, 43^

We found that plasma GFAP levels were higher in CU females than males, consistent with prior studies.^44, 45^ In sex-stratified analyses, GFAP’s association with cognitive decline and clinical progression was significant mainly in females. Although baseline cortical volume in AD-signature regions did not differ by sex, higher GFAP predicted greater longitudinal atrophy only in females. Associations with amyloid burden did not vary by sex, consistent with evidence that amyloid burden itself does not differ between males and females at any age.^46–48^ However, in Aβ-participants from the LEARN study, higher baseline GFAP levels predicted future amyloid positivity exclusively in females. These results suggest that astrocytic activation contributes more significantly to neurodegeneration and cognitive decline in females, highlighting GFAP’s potential as a sex-sensitive early biomarker and supporting the role of heightened neuroinflammation in female AD risk and progression.

Plasma GFAP increases with age,^17, 44^ and older individuals in our study had higher baseline levels. Despite this, associations with cognitive decline and clinical progression were observed in both age groups. In Aβ-participants from the LEARN cohort, GFAP’s link to baseline PACC and progression to CDR > 0 was stronger in younger individuals, suggesting that elevated GFAP may reflect early astrocytic activation from non-amyloid processes or a more aggressive pathology in younger people, predisposing them to decline even before significant amyloid accumulation.

A few limitations should be acknowledged. First, although some studies have demonstrated a direct association, there is no definitive proof that blood GFAP originates from reactive astrocytes in the brain.^49–51^ Second, GFAP is not specific to AD and may be elevated in other neurological or systemic conditions.^52, 53^ Finally, we analyzed only baseline GFAP; longitudinal trajectories may better capture temporal dynamics across disease stages.

In conclusion, plasma GFAP holds promise as a prognostic blood-based biomarker for predicting cognitive decline, risk of clinical progression, amyloid burden, and AD-related brain atrophy, with stronger effects in females. These findings highlight GFAP’s potential utility for early risk stratification and sex-specific applications. Future work should validate these results in multi-cohort studies and assess whether GFAP can improve trial enrichment and guide preventive interventions.

## Author Contributions

Elham Ghanbarian (Conceptualization; Methodology; Formal analysis; Writing - Original Draft; Investigation); Tianchen Qian (Data analysis, Methodology, Writing - Review & Editing); Babak Khorsand (Writing - Review & Editing); Lukai Zheng (Writing - Review & Editing), S. Ahmad Sajjadi (Writing - Review & Editing); Joshua Grill (Writing - Review & Editing), Laura Rabin (Conceptualization; ; Writing - Review & Editing); Richard Lipton (Conceptualization; Writing - Review & Editing); Reisa Sperling (Writing - Review & Editing), Ali Ezzati (Conceptualization; Methodology; Investigation; Writing - Review & Editing).

## Acknowledgements

This study was supported in part by grants from the National Institute of Health (NIA K23 AG063993; R01 AG080635; R01AG095017); the Alzheimer’s Association (SG-24-988292 ISAVRAD); Cure Alzheimer’s Fund.

## Source of data and Funding

The A4 Study is a secondary prevention trial in preclinical Alzheimer’s disease, aiming to slow cognitive decline associated with brain amyloid accumulation in clinically normal older individuals. The A4 Study is funded by a public-private philanthropic partnership, including funding from the National Institutes of Health-National Institute on Aging (U19 AG010483, U24AG057437, R01 AG063689), Eli Lilly and Company, Alzheimer’s Association, Accelerating Medicines Partnership, GHR Foundation, an anonymous foundation and additional private donors, with in-kind support from Avid and Cogstate. The companion observational Longitudinal Evaluation of Amyloid Risk and Neurodegeneration (LEARN) Study is funded by the Alzheimer’s Association (LEARN-15-338729) and GHR Foundation. The A4 and LEARN Studies are led by Dr. Reisa Sperling at Brigham and Women’s Hospital, Harvard Medical School and Dr. Paul Aisen at the Alzheimer’s Therapeutic Research Institute (ATRI), University of Southern California. The A4 and LEARN Studies are coordinated by ATRI at the University of Southern California, and the data are made available through the Laboratory for Neuro Imaging at the University of Southern California. The participants screening for the A4 Study provided permission to share their de-identified data to advance the quest to find a successful treatment for Alzheimer’s disease.

## Data availability

The data that support the findings of this study is openly available in https://www.a4studydata.org/.

## Supplementary Material

**Supplementary Table S1.** Sample sizes for imaging and plasma biomarkers by study cohort.

|  | Total | A4-Placebo | A4-Solanezumab | LEARN |
| --- | --- | --- | --- | --- |
| **Amyloid PET** |  |  |  |  |
| Cross-sectional | 949 | 245 | 254 | 450 |
| Longitudinal | 421 | 175 | 0 | 246 |
| **MRI** |  |  |  |  |
| Cross-sectional | 494 | 241 | 253 | 0 |
| Longitudinal | 172 | 172 | 0 | 0 |
| **Plasma p-tau217** |  |  |  |  |
| Cross-sectional | 695 | 213 | 216 | 266 |
| Longitudinal | 162 | 162 | 0 | 0 |

Abbreviations: p-tau217, phosphorylated tau at threonine 217

**Supplementary Table S2.** Association of baseline GFAP with baseline PACC scores in the entire sample and stratified by age and sex.

|  | Total (n=695) |  |  | Female (n=418) |  |  | Male (n=277) |  |  |
| --- | --- | --- | --- | --- | --- | --- | --- | --- | --- |
| | $\beta$ | SE | p value | $\beta$ | SE | p value | $\beta$ | SE | p value |
| Entire sample | -4.34 | 1.52 | 0.004 | -5.31 | 1.93 | 0.006 | -2.67 | 2.48 | 0.283 |
| Age 65-75 (n=573) | -5.80 | 1.89 | 0.002 | -6.27 | 2.24 | 0.005 | -4.42 | 3.54 | 0.213 |
| Age > 75 (n=122) | -7.49 | 2.68 | 0.006 | -8.59 | 4.02 | 0.037 | -6.58 | 3.43 | 0.06 |

Entire sample n=695, Age 65-75: n=573 (n=351 female, n=222 male), Age > 75 n=122 (n=67 female, n=55 male)

**Supplementary Table S3.** Association of baseline plasma GFAP with the rate of change in PACC scores in the entire sample and stratified by age and sex.

|  | Total (n=695) |  |  | Female (n=418) |  |  | Male (n=277) |  |  |
| --- | --- | --- | --- | --- | --- | --- | --- | --- | --- |
| | $\beta$ | SE | p value | $\beta$ | SE | p value | $\beta$ | SE | p value |
| Entire sample | -0.17 | 0.03 | <0.001 | -0.21 | 0.04 | <0.001 | -0.14 | 0.06 | 0.017 |
| Age 65-75 (n=573) | -0.10 | 0.04 | 0.018 | -0.17 | 0.05 | 0.002 | -0.006 | 0.08 | 0.931 |
| Age > 75 (n=122) | -0.21 | 0.08 | 0.010 | -0.16 | 0.12 | 0.202 | -0.22 | 0.1 | 0.034 |

Entire sample n=695, Age 65-75: n=573 (n=351 female, n=222 male), Age > 75 n=122 (n=67 female, n=55 male)

**Supplementary Table S4.** Associations of baseline plasma GFAP with risk of progression to CDR > 0 in the entire sample and stratified by age and sex.

| Groups | Total (n=659) |  | Female (n= 392) |  | Male (n= 267) |  |
| --- | --- | --- | --- | --- | --- | --- |
|  | Hazard Ratio | p value | Hazard Ratio | p value | Hazard Ratio | p value |
| Entire sample | 1.24 | 0.002 | 1.34 | 0.003 | 1.16 | .119 |
| Age 65-75 (n=545) | 1.31 | 0.006 | 1.30 | 0.045 | 1.33 | .063 |
| Age > 75 (n=114) | 1.26 | 0.015 | 1.48 | 0.009 | 1.14 | .296 |

Entire sample n=659, Age 65-75: n=545 (n=331 female, n=214 male), Age > 75 n=114 (n=61 female, n=53 male)

